# Non-linear relationship between serum phosphate and 30-day mortality in critically ill patients with cardiogenic shock: a cohort study from MIMIC-IV

**DOI:** 10.1101/2025.05.20.25327987

**Authors:** Shuyuan Qi, Yingxiu Huang, Shuying Qi, Guangyao Zhai

## Abstract

**Objective:** Cardiogenic shock (CS) carries high mortality despite management advances. Elevated phosphate links to poor outcomes in critical illness, but its role in CS remains unclear. This study explored the association between serum phosphate and 30-day mortality in CS patients.

**Methods:** A retrospective cohort study using the MIMIC-IV database stratified 2308 CS patients by serum phosphate tertiles (T1: <3.9 mg/dL, T2: 3.9–5.3 mg/dL, T3: ≥5.3 mg/dL) measured ≤24h post-ICU admission. Cox regression analyzed associations between phosphate levels and 30-day (primary outcome) and in-hospital mortality (secondary outcome). Kaplan-Meier curves and restricted cubic spline analysis were conducted to evaluate survival disparities and non-linear relationships.

**Results:** The 30-day and in-hospital mortality rates were 39.3% and 35.8%, respectively. Phosphate elevation independently correlated with heightened mortality after multivariable adjustment (adjusted hazard ratios [HR] 1.08, 95% CI 1.03–1.13, p = 0.001 for both 30-day and in-hospital mortality). Mortality risk escalated across phosphate tertiles (T2 vs. T1: HR 1.43, 95% CI 1.16–1.75, p = 0.001; T3 vs. T1: HR 1.70, 95% CI 1.34–2.16, p < 0.001). A non-linear inflection point occurred at 5.52 mg/dL: each 1 mg/dL increase below this threshold raised 30-day mortality risk by 17% (adjusted HR 1.17, 95% CI 1.04–1.32, p = 0.0087), with risk plateauing above it. Kaplan-Meier curves confirmed significant survival differences (p < 0.001).

**Conclusion:** Elevated serum phosphate levels are independently associated with increased both 30-day and in-hospital mortality in CS patients, with a nonlinear inflection point at 5.5 mg/dL. Serum phosphate may serve as a potential biomarker for risk assessment and therapeutic guidance in CS. Prospective validation is warranted.

## Introduction

Cardiogenic shock (CS) is a clinical syndrome due to reduced cardiac output, characterized by systemic hypoperfusion with inadequate tissue perfusion[1]. CS-related hospitalizations have increased steadily from 122 per 100,000 in 2004 to 408 per 100,000 in 2018[2]. In the past, acute myocardial infarction (AMI) was the predominant etiology of CS[3]. Nevertheless, recent data from a United States registry indicated that only approximately one-third of CS admissions were directly attributable to AMI, while the remaining two-thirds of CS cases stemmed from a variety of other etiologies, such as ischemic cardiomyopathy (ICM), non-ICM, and valvular heart diseases[4]. Despite advancements in treatment strategies, the short-term mortality rate for CS remains as high as 30%∼50%[3,5–9]. Given the severity of CS, it imposes a substantial economic burden, with a median total 1-year cost of $45,713 for those surviving to discharge[10]. The prognosis of patients with CS is multifactorial, involving baseline clinical profiles, disease severity stratification, cardiac and non-cardiac comorbidities, and time-sensitive therapeutic optimization. Evidence-based predictors of mortality encompass but is not confined to, advanced age, sustained hypotension, escalating inotropic requirements, persistent lactate elevation, acute kidney injury, pre-existing heart failure, and neurological impairment[11–14]. However, current investigations into CS risk stratification remain limited, posing a challenge to clinical practice. Identifying validated predictors is imperative for prognostic stratification, thereby underpinning risk-tailored therapies.

Phosphate, as an essential mineral component in the human body, is involved in multiple physiological processes, including energy metabolism, bone mineralization, acid–base balance regulation, and cellular signal transduction[15]. The kidneys primarily regulate phosphate homeostasis[15]. Numerous studies have confirmed that elevated serum phosphate levels are linked to increased mortality in various diseases, such as acute myocardial infarction[16], chronic kidney disease[17,18], chronic obstructive pulmonary disease (COPD)[19], septic shock[20], ischemic stroke[21], blunt trauma[22] and acute pancreatitis[23]. Pooled evidence from a meta-analysis has established a mortality risk elevation linked to hyperphosphatemia among intensive care populations[24]. However, none of previous studies specifically focused on CS patients, leaving the prognostic value of phosphate imbalance in this cohort unclear.

To address this gap, this study is designed to investigate whether serum phosphate concentrations within 24 hours of ICU admission correlate with 30-day mortality risk among CS patients in the intensive care unit (ICU). We hypothesize that admission serum phosphate levels are associated with 30-day mortality in CS patients.

## Materials and Methods

### Data sources

We analyzed data from the Medical Information Mart for Intensive Care (MIMIC)-IV[25] (version 3.1), a deidentified critical care database containing high-fidelity electronic health records of ICU patients at Beth Israel Deaconess Medical Center (BIDMC) in Boston, Massachusetts, USA (2008-2022). Ethical approval including informed consent waiver was granted by Massachusetts Institute of Technology (MIT) and BIDMC Institutional Review Boards. Shuyuan Qi (Certification No.14251403) and Yingxiu Huang (Certification No.56513391) have completed an online training program on human research protection, sponsored by US National Institutes of Health. The MIMIC-IV database was accessed for research purposes via PhysioNet from March 6 to March 13, 2025. Throughout data collection and subsequent analysis, researchers had no access to any identifiable patient information, ensuring complete anonymity. This research adhered to the Strengthening the Reporting of Observational Studies in Epidemiology (STROBE) reporting guideline[26].

### Study population

The MIMIC-IV database initially provided 94,458 eligible ICU patients, from which 29,092 individuals were excluded on the basis of repeated hospital stays and ICU visits. Patients identified with CS from 2008 to 2022 were included in this cohort. The diagnosis was verified utilizing the International Classification of Diseases (ICD) codes from the Ninth and Tenth revisions (785.51, R57.0, T81.11XA). All CS participants were aged 18 years or older. Only the initial hospital admission and the initial ICU admission for each patient were selected for analysis (n=2,441). Exclusion criteria included absence of phosphate measurements within 24 hours of ICU admission (n=127) and presence of phosphate level outliers (n=6) (Fig.1).

**Fig 1.**
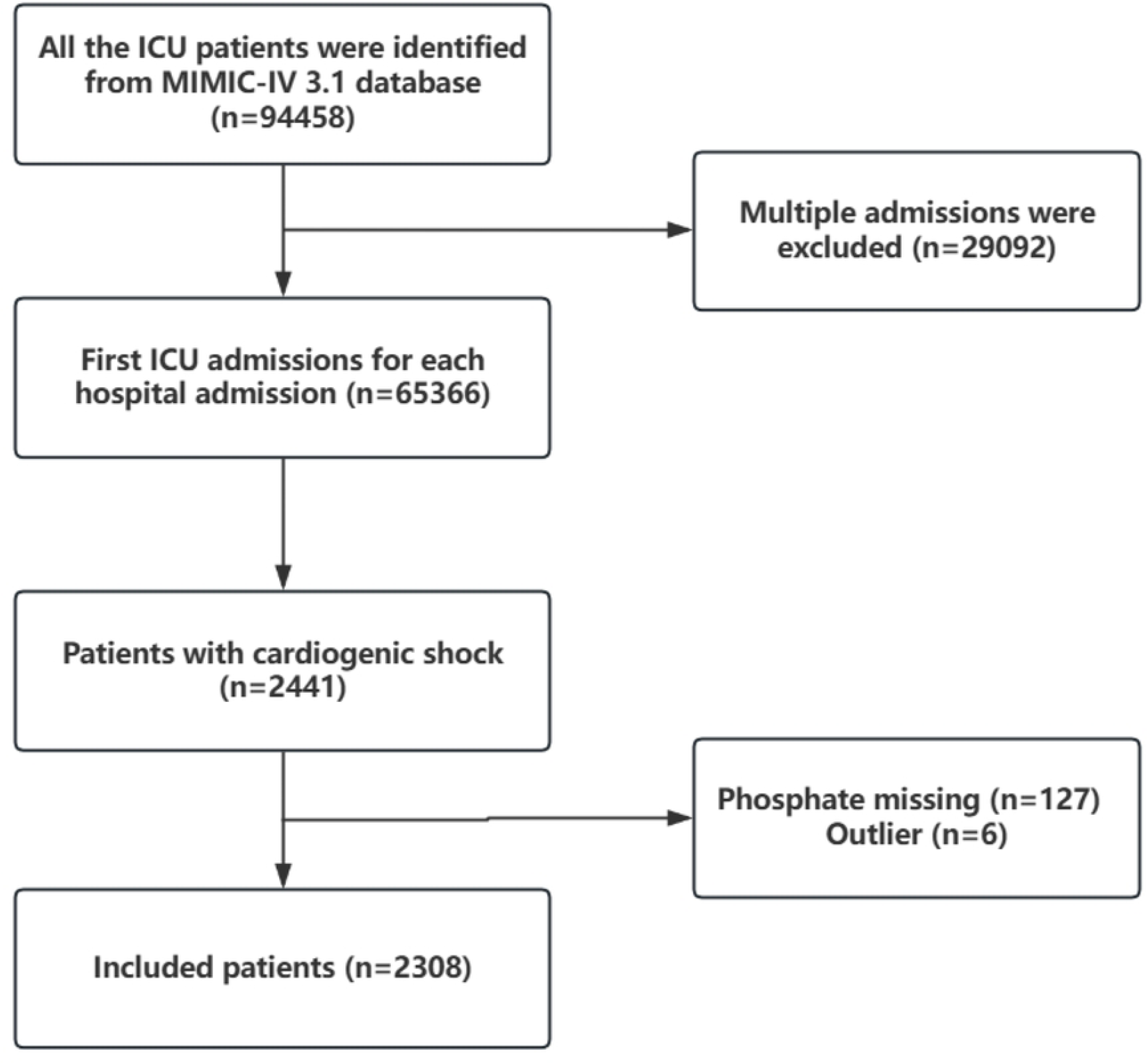
The flow chart of the study. The final cohort comprised 2,308 patients after exclusion of those with: (1) repeated hospital admissions and ICU readmissions; (2) missing or outlier serum phosphate measurements.

### Dataset derivation

Using Structured Query Language (SQL), we extracted MIMIC-IV data into PostgreSQL database for further analysis. The dataset encompasses patient demographics (age, sex, ethnicity, height, weight), physiological parameters (respiratory and heart rates, mean arterial pressure [MAP], and body temperature), comorbid conditions (congestive heart failure, hypertension, AMI, rheumatic disease, cerebrovascular disease, dementia, chronic pulmonary disease, peripheral vascular disease, peptic ulcer disease, diabetes mellitus with and without chronic complications [cc], renal disease, malignant cancer, severe liver disease, sepsis, paraplegia) and disease seriousness (acute physiology score III [APSIII], Charlson comorbidity index [CCI]). Comorbidity data were obtained using the ICD coding system. Intervention measures were also recorded, including intra-aortic balloon pumping (IABP), percutaneous coronary intervention (PCI), coronary artery bypass grafting (CABG), ventilation (non-invasive ventilation and invasive ventilation), vasoactive agents, and renal replacement therapy (RRT). Additionally, the dataset included the laboratory measurements from the initial day of ICU stay, such as phosphate, glucose, hemoglobin, white blood cell (WBC), platelet, anion gap, blood urea nitrogen (BUN), calcium, chloride, creatinine, sodium, potassium, partial thromboplastin time (PTT), alanine aminotransferase (ALT), creatine kinase-myocardial Band (CKMB) and lactate. The calculation of body mass index (BMI, kg/m^2^) was weight (kg) divided by height squared (m^2^). The estimated glomerular filtration rate (eGFR) was calculated using the 2021 Chronic Kidney Disease Epidemiology Collaboration (CKD-EPI) equation[27]. Outcomes of the patients were also documented.

### Exposure and outcome

The primary exposure factor in this study was serum phosphate level, assessed as a continuous variable. In addition, serum phosphate level was categorized into tertiles for analysis: T1 (<3.9 mg/dL), T2 (3.9–5.3 mg/dL), and T3 (≥5.3 mg/dL). The baseline phosphate concentration was determined based on the maximum measurement within the first 24 hours of ICU admission. The follow-up period was at least 30 days. The primary outcome was 30-day mortality. In-hospital mortality was the secondary outcome.

### Statistical analysis

Descriptive statistics were employed to analyze patient characteristics. Continuous variables were expressed as mean ± standard deviation (SD) or median (interquartile range [IQR]) based on the distribution of the data. Categorical variables were presented as frequencies and percentages. To assess the baseline characteristics, we employed the chi-square test for categorical variables. One-way ANOVA and Kruskal-Wallis tests were implemented for normally and non-normally distributed continuous variables, respectively. Confounders were selected based on clinical significance and variables with p value less than 0.05 in the univariate analysis. Using Cox proportional hazards regression, we constructed five models to assess the association between serum phosphate (continuous and tri-categorized) and mortality outcomes, expressed as hazard ratios (HR) with 95% confidence intervals (CI). Model 1 was crude model without adjustment. Model 2 adjusted for age, sex, race and BMI. Model 3 further adjusted for respiratory and heart rates, MAP, temperature, hemoglobin, WBC, anion gap, BUN, calcium, chloride, potassium, PTT, lactate and eGFR. Model 4 included adjustments for peripheral vascular disease, heart failure, dementia, cerebrovascular disease, diabetes with cc, malignant cancer, renal disease, sepsis, CCI, APSIII and model 3. Model 5 designated as the primary model, incorporated adjustments for IABP, CABG, ventilation, vasoactive agents, RRT and Model 4. Curve fitting methods were utilized to investigate the phosphate-mortality associations. Restricted cubic splines (knots at 5th/35th/65th/95th percentiles) were employed. Additionally, survival outcomes were compared using Kaplan-Meier curves stratified by serum phosphate tertiles. We performed stratified analyses to assess potential interaction effects by age, sex, eGFR, AMI and congestive heart failure. The maximum proportion of missing values across variables was 35.4%. To address missing data, we implemented a multivariate single imputation method based on an iterative imputer, using a Bayesian Ridge model as the estimator at each step of the round-robin imputation process. A single imputed dataset was generated and used for primary analyses. We also conducted sensitivity analyses via complete-case analysis (excluding cases with missing data) to evaluate result robustness. All analyses were performed using R version 4.2.2 and the Free Statistics software version 2.1 was employed. A two-sided p-value of less than 0.05 was considered statistically significant.

## Results

### Baseline characteristics of patients

A total of 2,308 CS participants were enrolled. The cohort had an average age of 68.1 years, with 59.4% being male. Baseline characteristics of patients categorized by serum phosphorus tertiles were presented in Table 1. Significant intergroup differences were observed in heart rate (p=0.041), MAP, respiratory rate and temperature (all p < 0.001). Group T3 exhibited elevated levels of glucose, WBC, anion gap, BUN, potassium, creatinine, PTT, ALT and lactate (all p < 0.001). Conversely, lower levels of hemoglobin, calcium, chloride, sodium, and eGFR were presented in group T3 (all p < 0.001). Group T3 showed the highest prevalence of comorbidities, including severe liver disease, renal disease, sepsis, diabetes with cc (all p < 0.001) and congestive heart failure (p = 0.008). Illness severity was greater in T3, reflected by higher APSIII scores and CCI (p < 0.001). Fewer patients underwent PCI and CABG while interventions such as ventilation, vasoactive agents and RRT were more frequent in group T3 (all p < 0.001). No significant differences were observed in gender, age, BMI or baseline comorbidities like hypertension and AMI (all p > 0.05).

**Table 1.**
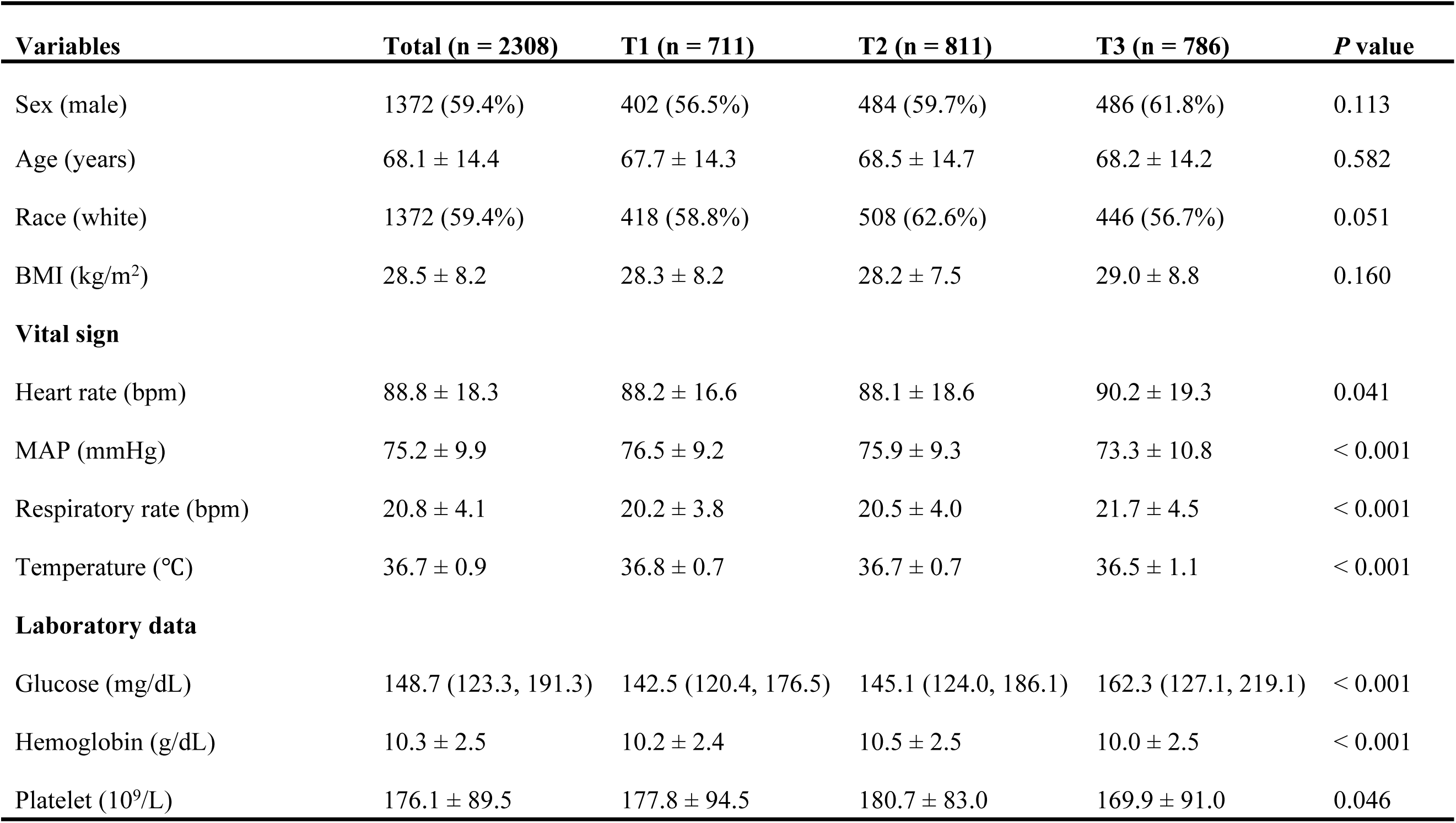
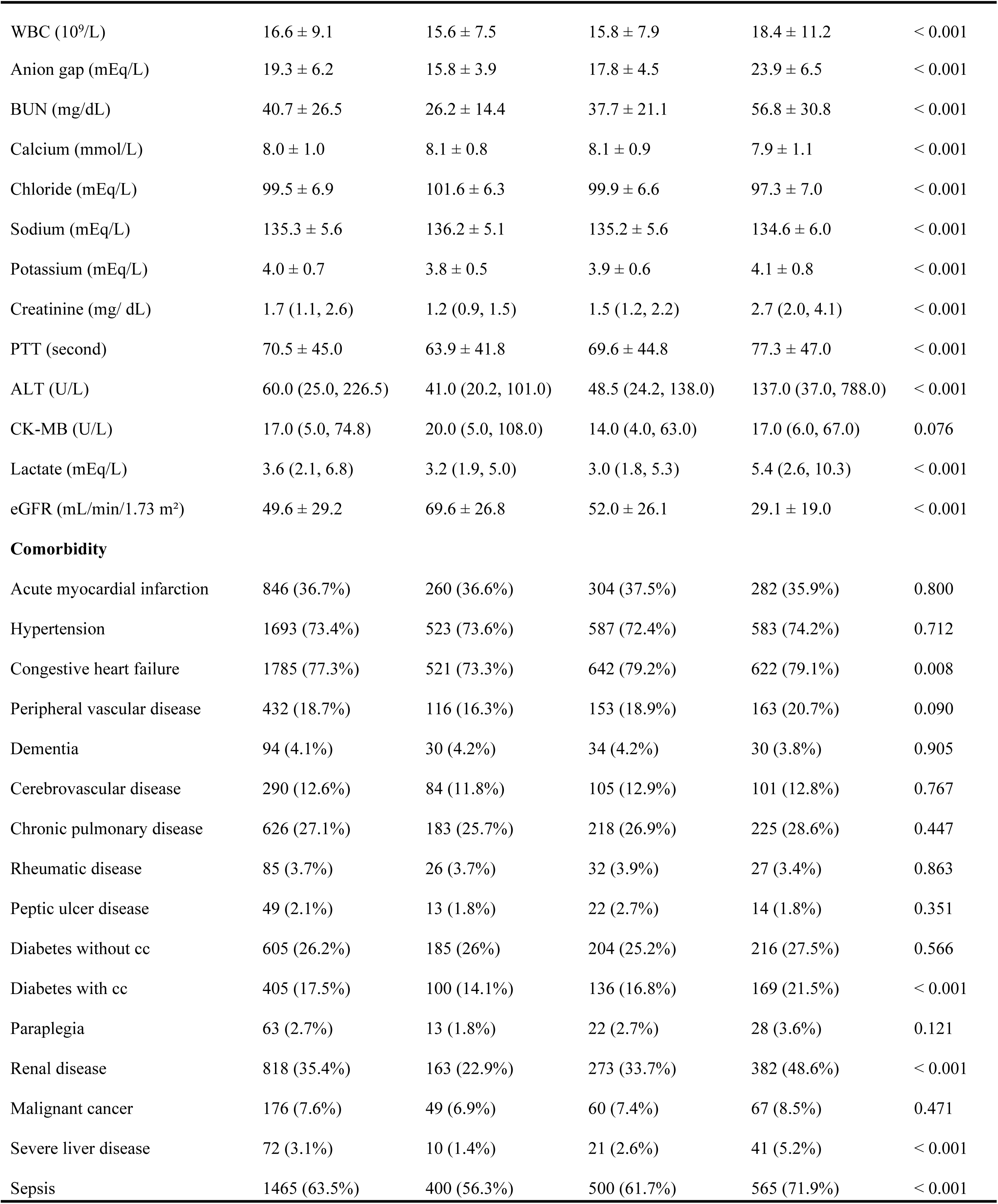
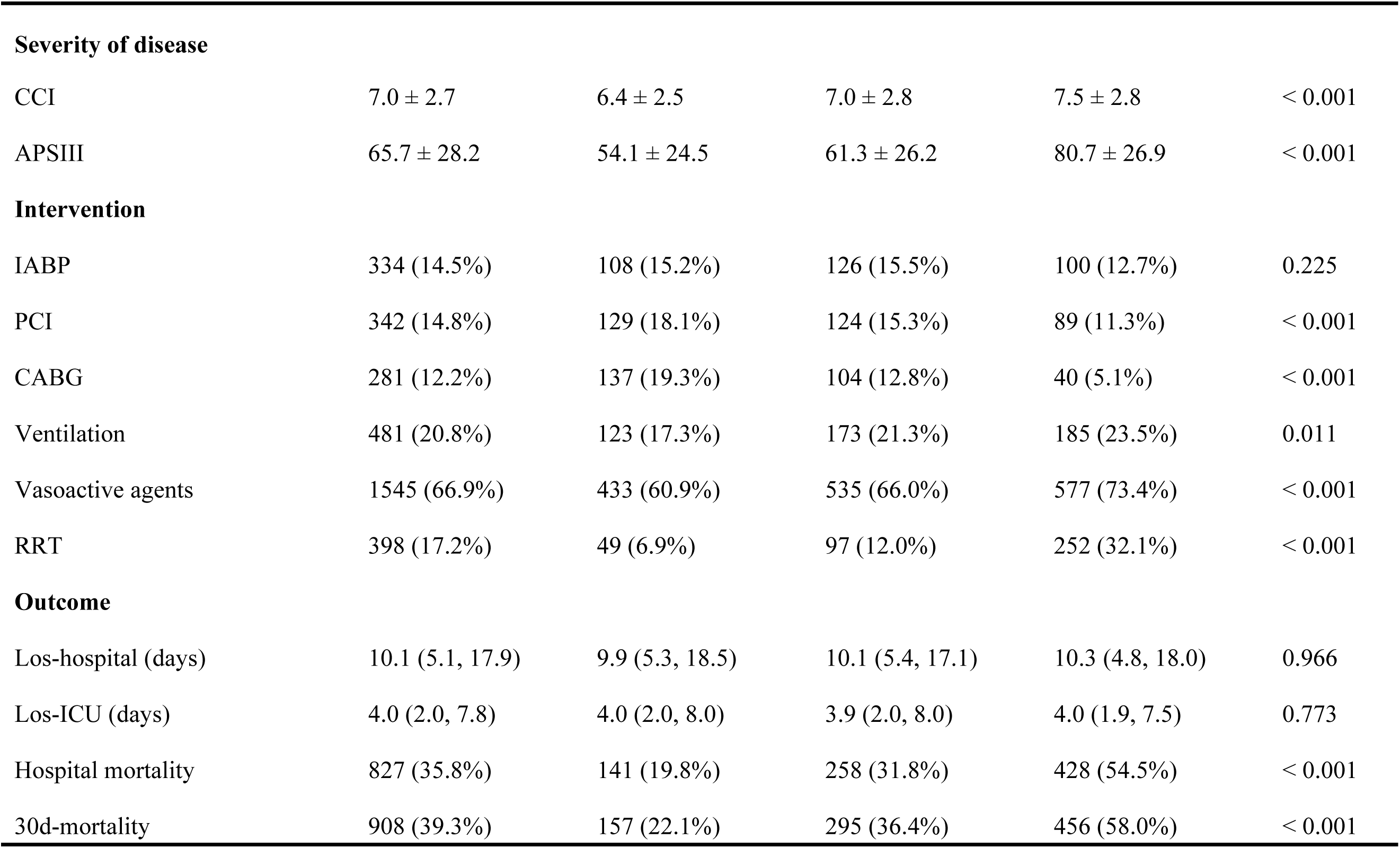
Study population characteristics at baseline.

### Association of serum phosphate levels with the mortality **outcomes**

The 30-day all-cause mortality rate was 39.3%, with in-hospital mortality at 35.8%. Group T3 exhibited a notably higher 30-day death rate (58.0%) than T1 (22.1%) and T2 (36.4%) (p < 0.001). Similar trends were observed for in-hospital mortality.

Univariate analysis identified multiple mortality-associated factors across demographic (age, sex, race), clinical (BMI, vital signs), laboratory (WBC, hemoglobin, lactate, eGFR, anion gap, BUN, electrolytes), comorbidities (heart failure, diabetes, peripheral vascular disease, renal disease, cerebrovascular disease, dementia, malignant cancer and sepsis), disease severity (CCI, APSIII) and treatment-related (IABP, CABG, RRT, vasoactive agents and ventilation) variables for both in-hospital and 30-day mortality (S1 Table).

When analyzed as a continuous variable, each unit increase in serum phosphate concentration was associated with a 31% elevated risk of 30-day mortality (crude HR 1.31, 95% CI: 1.28–1.35) and a 32% increased risk of in-hospital mortality (crude HR 1.32, 95% CI: 1.28–1.36) in unadjusted analyses, both with p < 0.001. After adjusting for all confounding factors in the multivariate Cox regression analysis, the hazard ratios of serum phosphate were identical for both 30-day mortality (HR 1.08, 95% CI: 1.03– 1.13, p = 0.001) and in-hospital mortality (HR 1.08, 95% CI: 1.03–1.13, p = 0.001). Per-unit (1 mg/dL) phosphate increment conferred 8% greater mortality risk for both endpoints. When phosphate levels were categorized, adjusted HRs for 30-day death rates in T2 (3.9–5.3 mg/dL) and T3 (≥5.3 mg/dL), compared with the lowest group (T1, <3.9 mg/dL), were 1.43 (95% CI: 1.16–1.75, p = 0.001) and 1.70 (95% CI: 1.34–2.16, p < 0.001), respectively, in the fully adjusted model. Similarly, higher phosphate tertiles showed progressively increased in-hospital mortality, with HRs of 1.40 (95% CI: 1.12– 1.73, p = 0.003) for T2 and 1.78 (95% CI: 1.39–2.27, p < 0.001) for T3 (Table 2).

**Table 2.** COX regression models to assess the association between serum phosphate levels and mortality outcomes.

|  | Variable (mg/dL) | Model 1 |  | Model 2 |  | Model 3 |  | Model 4 |  | Model 5 |  |
| --- | --- | --- | --- | --- | --- | --- | --- | --- | --- | --- | --- |
|  |  | HR (95%CI) | P | HR (95%CI) | P | HR (95%CI) | P | HR (95%CI) | P | HR (95%CI) | P |
| **30-day mortality** | Phosphate | 1.31 (1.28~1.35) | <0.001 | 1.33 (1.3~1.37) | <0.001 | 1.10 (1.05~1.15) | <0.001 | 1.10 (1.05~1.15) | <0.001 | 1.08 (1.03~1.13) | 0.001 |
|  | Phosphate tertile |  |  |  |  |  |  |  |  |  |  |
|  | T1 (<3.9) | 1(Ref) |  | 1(Ref) |  | 1(Ref) |  | 1(Ref) |  | 1(Ref) |  |
|  | T2 (3.9~5.3) | 1.79 (1.48~2.18) | <0.001 | 1.78 (1.47~2.17) | <0.001 | 1.51 (1.23~1.85) | <0.001 | 1.48 (1.21~1.82) | <0.001 | 1.43 (1.16~1.75) | 0.001 |
|  | T3 (≥5.3) | 3.74 (3.12~4.49) | <0.001 | 3.83 (3.19~4.60) | <0.001 | 1.93 (1.53~2.44) | <0.001 | 1.81 (1.43~2.29) | <0.001 | 1.70 (1.34~2.16) | <0.001 |
|  | P for trend |  | <0.001 |  | <0.001 |  | <0.001 |  | <0.001 |  | <0.001 |
| **In-hospital mortality** | Phosphate | 1.32 (1.28~1.36) | <0.001 | 1.34 (1.3~1.38) | <0.001 | 1.10 (1.05~1.15) | <0.001 | 1.10 (1.05~1.15) | <0.001 | 1.08 (1.03~1.13) | 0.001 |
|  | Phosphate tertile |  |  |  |  |  |  |  |  |  |  |
|  | T1 (<3.9) | 1(Ref) |  | 1(Ref) |  | 1(Ref) |  | 1(Ref) |  | 1(Ref) |  |
|  | T2 (3.9~5.3) | 1.74 (1.42~2.13) | <0.001 | 1.74 (1.41~2.13) | <0.001 | 1.48 (1.19~1.83) | <0.001 | 1.45 (1.17~1.80) | 0.001 | 1.40 (1.12~1.73) | 0.003 |
|  | T3 (≥5.3) | 3.86 (3.19~4.67) | <0.001 | 3.93 (3.24~4.76) | <0.001 | 2.01 (1.57~2.57) | <0.001 | 1.89 (1.48~2.42) | <0.001 | 1.78 (1.39~2.27) | <0.001 |
|  | P for trend |  | <0.001 |  | <0.001 |  | <0.001 |  | <0.001 |  | <0.001 |

Note: HR-hazard ratio, CI-confidence interval; BMI-body mass index; MAP- mean arterial pressure; BUN-blood urea nitrogen; WBC-white blood cell; PTT-partial thromboplastin time; eGFR-estimated glomerular filtration rate; cc-chronic complication; CCI-Charlson comorbidity index; APS-acute physiology score; IABP-intra-aortic balloon pumping; CABG-coronary artery bypass grafting; RRT-renal replacement therapy.
Model 1: Unadjusted;
Model 2: Adjusted for age, sex, race and BMI;
Model 3: Model 2 plus heart rate, MAP, respiratory rate, temperature, hemoglobin, WBC, anion gap, BUN, calcium, chloride, potassium, PTT, lactate and eGFR;
Model 4: Model 3 plus congestive heart failure, peripheral vascular disease, dementia, cerebrovascular disease, diabetes with cc, renal disease, malignant cancer, sepsis, CCI and APSIII;
Model 5: Model 4 plus IABP, CABG, ventilation, vasoactive agents and RRT.

A nonlinear association between phosphate concentrations and mortality outcomes was identified through restricted cubic spline analysis (non-linearity p = 0.018 for 30- day mortality; p = 0.025 for in-hospital mortality (Fig. 2A and 2B). By integrating graphical analysis with clinical insights, we identified a significant threshold for serum phosphate levels at 5.52 mg/dL. Below this threshold, each 1 mg/dL increment in serum phosphate independently linked to higher death rates (30-day: adjusted HR 1.17, 95% CI: 1.04–1.32, p = 0.0087; in-hospital: adjusted HR 1.17, 95% CI: 1.03–1.32, p = 0.147).

**Fig 2.**
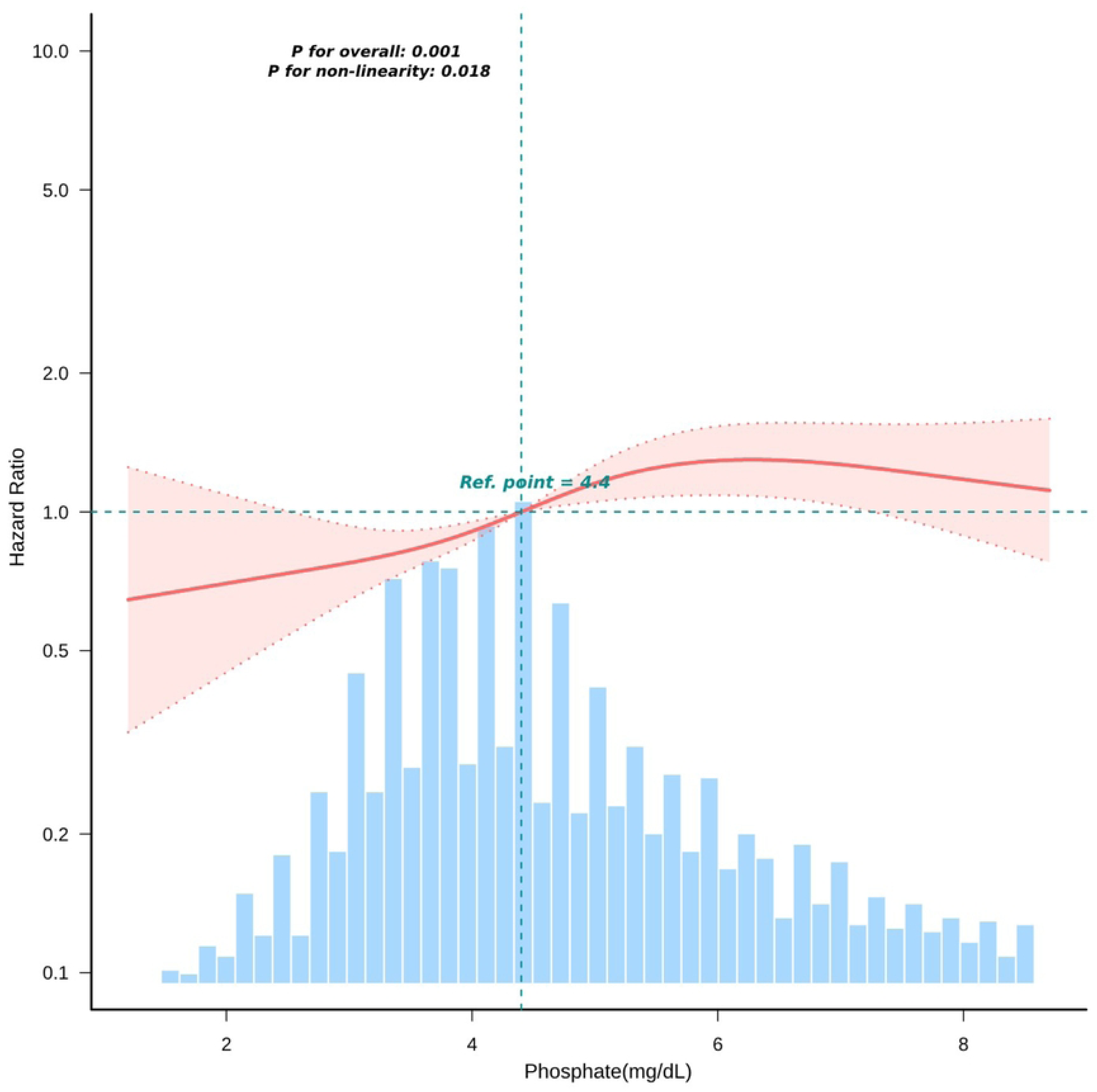
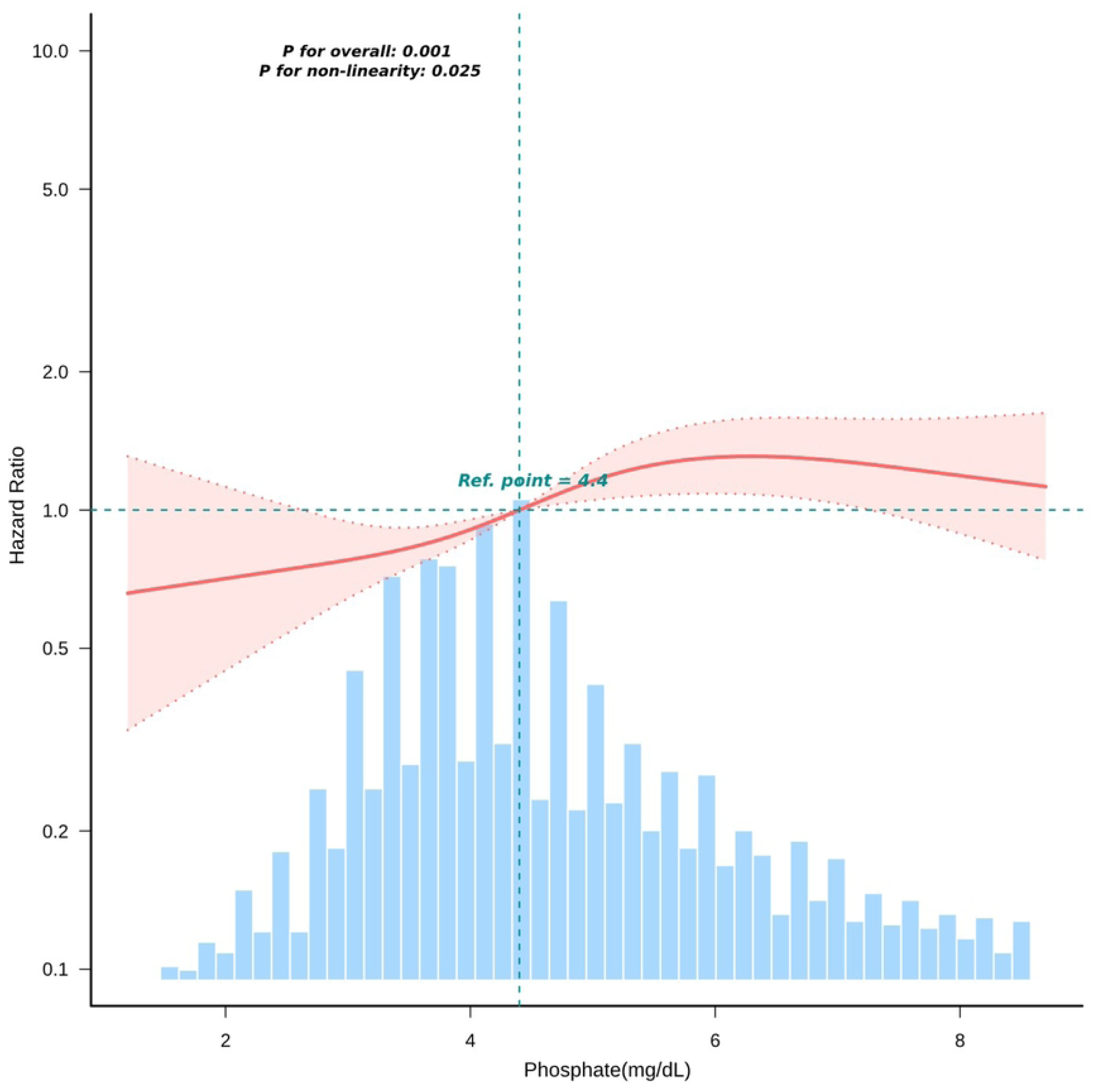
Nonlinear association between serum phosphate levels and mortality risks in cardiogenic shock patients. (A) Dose-response relationship between serum phosphate (mg/dL) and 30-day mortality by restricted cubic spline analysis (P for overall = 0.001, P for non-linearity = 0.018). (B) Corresponding in-hospital mortality analysis (P for non-linearity = 0.025), both adjusted for Model 5 covariates. Shaded areas represent 95% confidence intervals. Only 95% of the data is displayed.

Notably, this association attenuated above the threshold (30-day: HR=1.03, 95% CI: 0.94–1.13; p = 0.5066; in-hospital: HR=1.04, 95% CI:0.95–1.14; p = 0.417), suggesting a differential mortality risk profile across phosphate strata (Table 3).

**Table 3.** Threshold effect analysis of phosphate-mortality association.

| Serum phosphate (mg/dL) | 30-day Mortality |  | Hospital Mortality |  |
| --- | --- | --- | --- | --- |
|  | HR (95%CI) | *P* value | HR (95%CI) | *P* value |
| <5.52 mg/dL | 1.171 (1.041,1.318) | 0.0087 | 1.167 (1.031,1.321) | 0.0147 |
| ≥5.52 mg/dL | 1.031 (0.943,1.126) | 0.5066 | 1.038 (0.948,1.137) | 0.4170 |
| Likelihood Ratio test |  | 0.0110 |  | 0.0210 |

Note: Adjusted for all covariates in Model 5.

### Kaplan–Meier survival analysis

Kaplan-Meier survival curves revealed significant survival disparities among serum phosphate tertiles. Patients in the highest tertile (T3; ≥5.3 mg/dL) exhibited markedly lower 30-day and in-hospital survival probability compared to those in the lowest tertile (T1;<3.9 mg/dL) (log-rank test, both p < 0.0001) (Fig. 3A and 3B).

**Fig 3.**
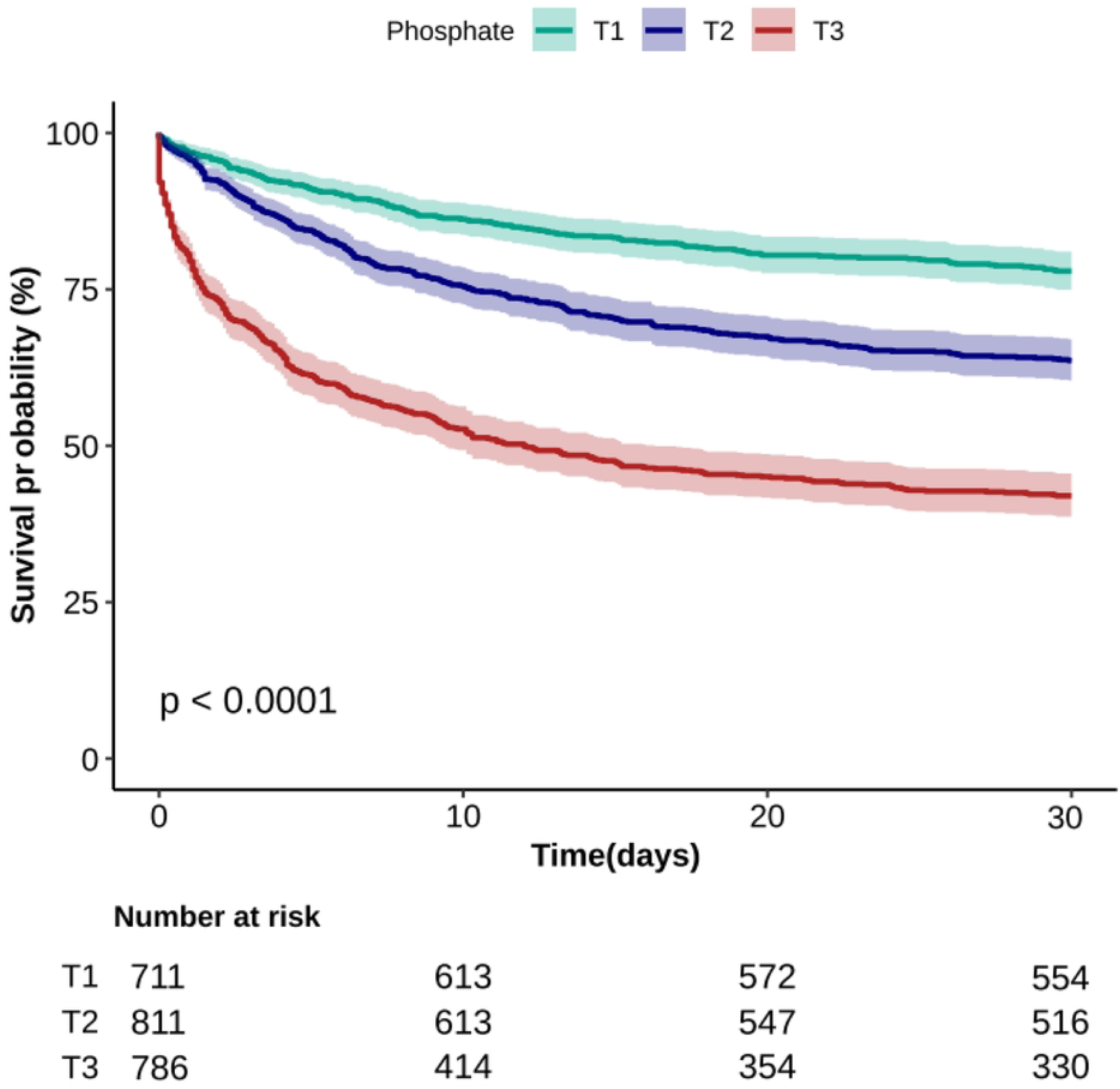
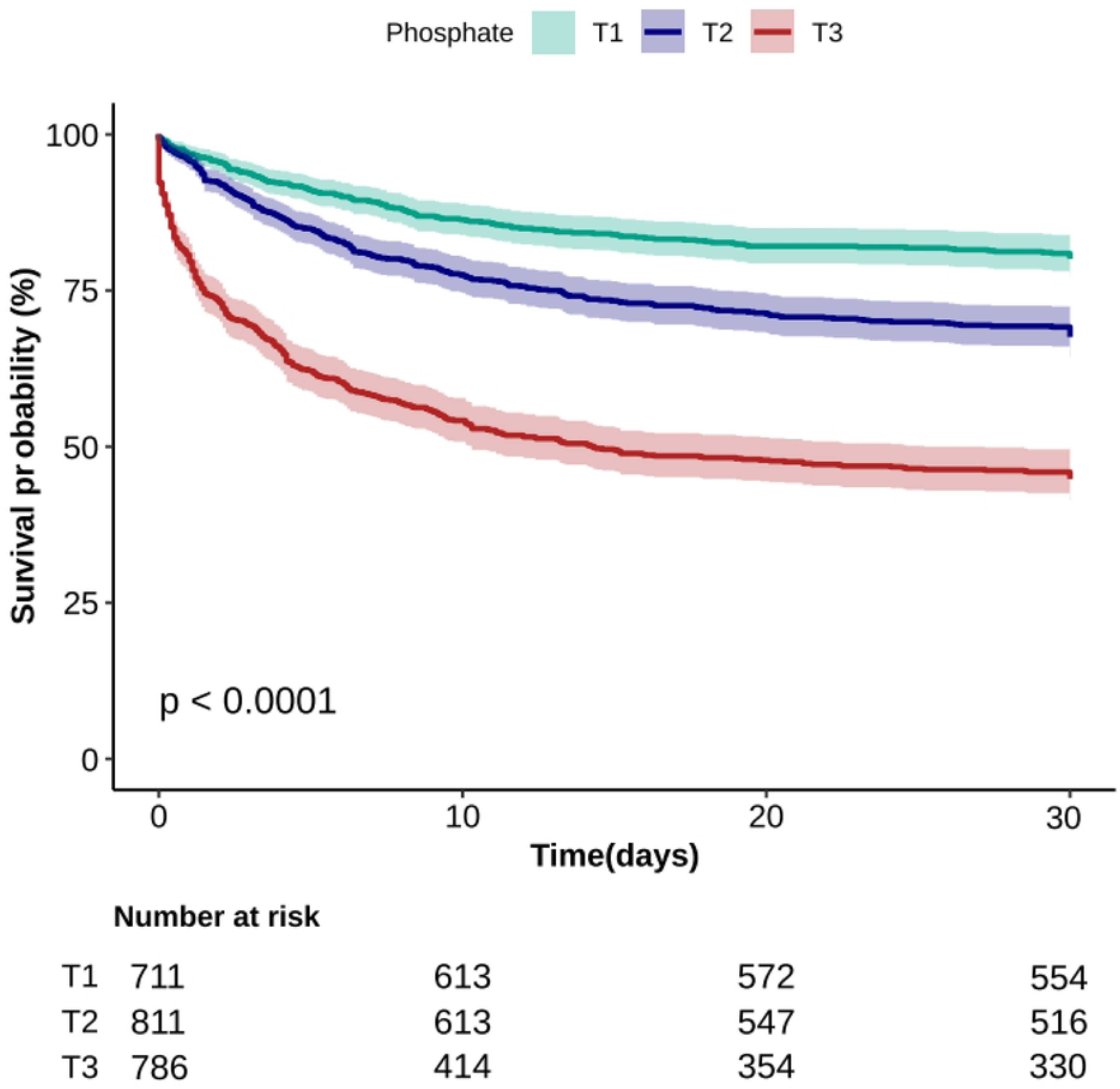
Kaplan-Meier survival analysis by serum phosphate tertiles in cardiogenic shock patients. (A) 30-day mortality and (B) in-hospital mortality curves are shown for patients categorized by phosphate tertiles. Log-rank tests demonstrated statistically significant differences across tertiles for both endpoints (30-day mortality: P<0.0001; in-hospital mortality: P<0.0001). Numbers at risk at baseline are displayed below each panel (T1=711, T2=811, T3=786).

### Subgroup analysis

Subgroup analyses revealed consistent mortality associations across predefined strata (sex, age, eGFR, AMI and congestive heart failure). While no interaction was found for 30-day mortality, an interaction between age groups (<65 years vs ≥65 years) and in-hospital mortality was observed (Fig. 4A and 4B).

**Fig 4.**
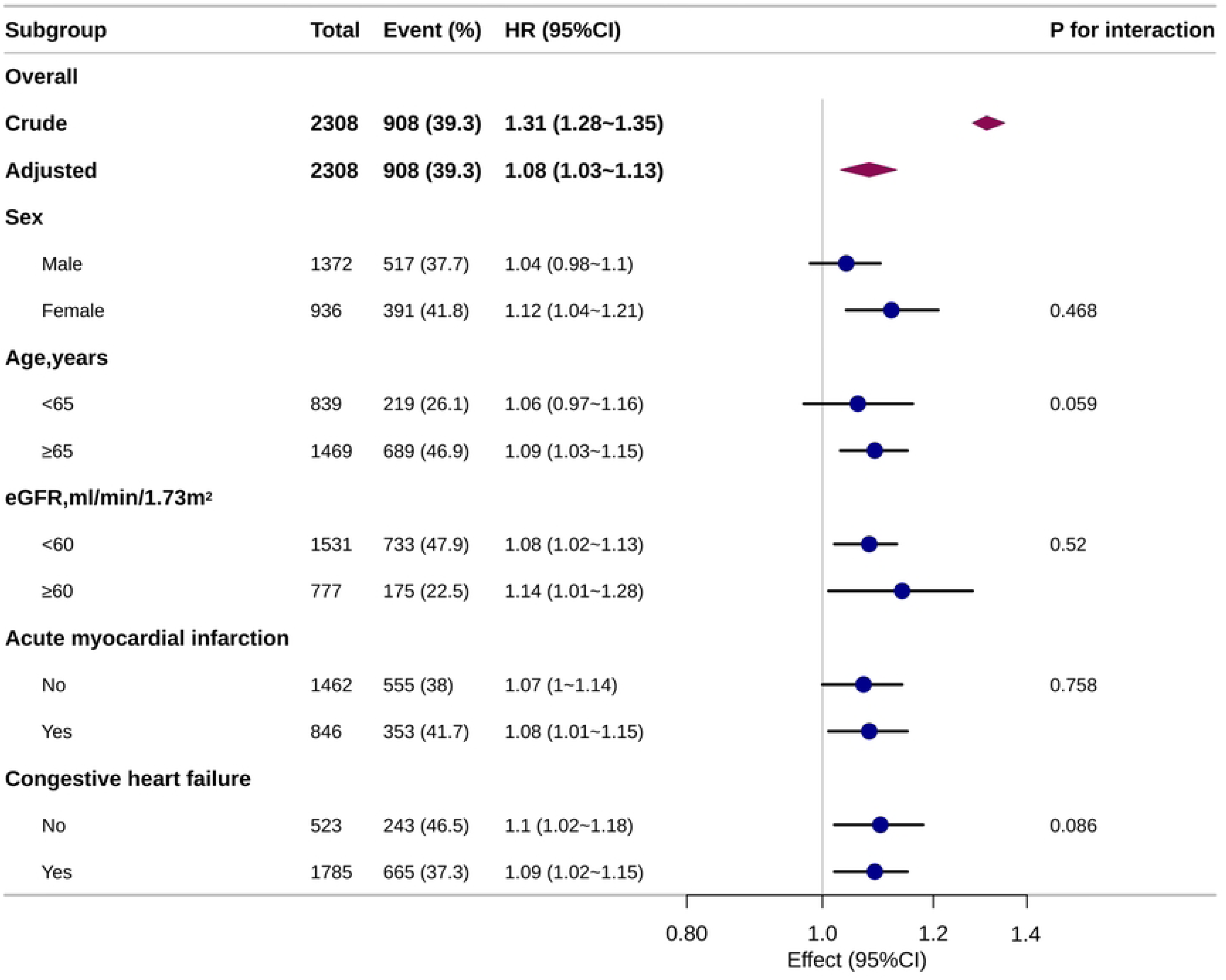
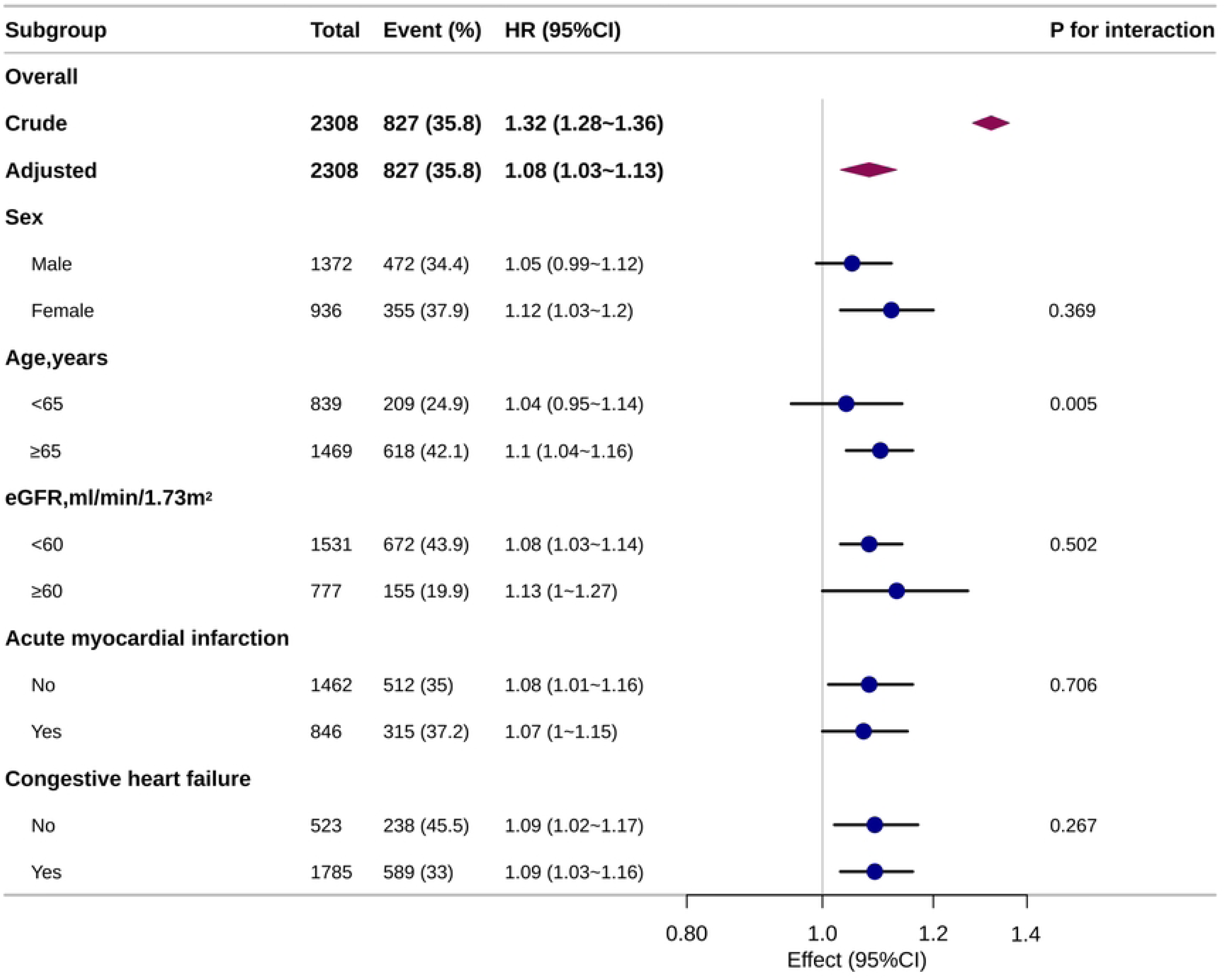
Forest plots of subgroup analyses for (A) 30-day mortality and (B) in-hospital mortality in cardiogenic shock patients: association with serum phosphate as a continuous variable. The forest plots display hazard ratios (HRs) with 95% confidence intervals for mortality outcomes across prespecified subgroups, adjusted for Model 5 covariates.

### Sensitivity analysis

We performed a complete-case analysis by excluding patients with missing values. Multivariate Cox regression analysis based on this subset yielded results consistent with primary analysis, confirming the robustness of our findings (S2 Table).

## Discussion

This study aimed to investigate the relationship between serum phosphate levels and mortality in critically ill patients with CS. Our analysis revealed a significant correlation between high serum phosphate concentrations and elevated mortality risks during hospitalization and within the 30-day follow-up period. Notably, we identified a non-linear relationship with a threshold point at 5.5 mg/dL: below this value, mortality risks progressively increased with rising phosphate levels (even within conventional normal ranges of 2.5-4.5 mg/dL), whereas risks plateaued beyond the threshold. These findings indicate that serum phosphate levels may offer novel insights for refined risk stratification and tailored therapeutic strategies in CS patients.

Phosphate imbalance is a common metabolic disorder in critically ill patients, with an overall incidence ranging from 17% to 45%[24]. Due to the high prevalence of myocardial dysfunction[28], activated inflammatory responses[29,30], and renal insufficiency[31] in patients with CS, impaired phosphate excretion and abnormal intracellular/extracellular distribution often lead to elevated serum phosphate levels. Our findings align with previous studies that have reported an association between increasing phosphate levels and mortality risk in critically ill patients[19,20,23,24,32]. However, our study extends this knowledge by demonstrating a non-linear relationship, which has not been previously described in the context of CS. Unlike other studies[20,21,32] that have shown a linear or U-shaped correlation between high phosphate levels and mortality risk in critically ill patients, we propose a novel "biphasic" pattern of phosphate-mortality association in CS: mortality risk increases linearly with phosphate levels below 5.5 mg/dL but plateaus beyond this threshold. This non-linear relationship suggests that the impact of phosphate levels on mortality in CS may be more complex than that in other diseases. The finding prompts reconsideration of the conventional reference range (2.5–4.5 mg/dL) and highlights the necessity for CS-specific phosphate management strategies, including revised targets and intensified monitoring protocols. Moreover, even subtle serum phosphate fluctuations within the upper normal range may independently influenced mortality outcomes, suggesting that CS patients exhibit lower tolerance to phosphate variability. Early detection of subclinical phosphate disorder and prompt stabilization of phosphate homeostasis hold promise for improving the prognosis in CS patients. Given its accessibility and cost-effectiveness, serum phosphate level may serve as a practical biomarker for prognostication in CS, particularly in resource-limited critical care settings.

The phosphate-mortality relationship observed in patients with CS may be explained by several potential mechanisms. Elevated serum phosphorus activates oxidative stress, characterized by excessive reactive oxygen species (ROS) generation and diminished endothelial nitric oxide synthase activity, resulting in endothelial dysfunction[33,34]. It is well-established that high phosphate levels drive mitochondrial dysfunction and induce a metabolic shift from oxidative phosphor ylation to glycolysis in cardiomyocytes, ultimately leading to energetic collapse and heart failure[33,35,36]. Also, high phosphate has been verified to induce osteogenic differentiation and calcification of vascular smooth muscle cells[37,38], thereby accelerating coronary artery calcification and arterial stiffness[39].Furthermore, serum phosphate elevation engages in pathological crosstalk with inflammation[29,30], through direct activation of pro-inflammatory pathways (nuclear factor kappa B, ROS), disruption of the fibroblast growth factor 23/Klotho axis, and promotion of a calcific microenvironment, which may exacerbate endothelial dysfunction, infiltrative myocardial inflammation, and calcific remodeling[40]. However, the precise mechanisms underlying this association remain to be fully elucidated, particularly the threshold effect of serum phosphate levels on mortality risk in CS. Further validation is needed.

Notably, after adjusting for eGFR and renal replacement therapy, elevated phosphate levels remained independently associated with mortality in CS patients. Subgroup analysis further revealed that the association persisted even in patients with eGFR ≥60 mL/min/1.73 m². Given prior evidence that phosphate may impair myocardial function through mechanisms involving endothelial dysfunction, oxidative stress, inflammation, and vascular calcification, we hypothesize that higher phosphate may exert direct cardiotoxic effects independent of renal function. However, further experimental verification is necessary.

The strength of this investigation lies in the use of the MIMIC-IV dataset, a multi- modal repository that integrates and standardizes large-scale cohorts, thereby enhancing the validity of the analysis. Subgroup analyses further confirm the robustness of the results. However, several limitations should be acknowledged. Firstly, the retrospective design may introduce selection bias. The sample was derived from a single medical center, limiting the generalizability of the findings. Secondly, our analysis relied on measurements of serum phosphate levels within the first 24 hours of ICU admission, which may not fully capture the dynamic changes in phosphate levels during the course of illness. Thirdly, many variables contained missing values due to data extraction from a public database. Although known confounders were adjusted for, residual confounding cannot be ruled out. Finally, as an observational study, we were unable to validate the hypothesized mechanisms linking phosphate levels to the severity and outcomes of CS. Future studies should aim to validate our findings in larger, more diverse patient populations. Multicenter, prospective studies with serial measurements of serum phosphate levels could provide insights into the dynamic relationship between phosphate levels and mortality in critically ill patients. Mechanistic studies are also needed to elucidate the biological pathways linking elevated phosphate levels to adverse outcomes in CS. Finally, interventional studies, such as randomized controlled trials of phosphate-binding agents, are needed to determine whether reducing serum phosphate levels can improve outcomes in critically ill patients with CS.

## Conclusion

The study highlights a non-linear relationship between early serum phosphate concentrations within 24 hours of ICU admission and mortality outcomes (in-hospital and 30-day), among patients with CS. A notable inflection point was detected at 5.5 mg/dL. Serum phosphate could serve as a potential biomarker for risk assessment and therapeutic guidance in ICU-admitted CS patients. Further research is required to validate these observations and elucidate the underlying mechanisms.

## Data availability statement

The data used in this study were obtained from the MIMIC-IV database (version 3.1, available at https://mimic.physionet.org/), which is publicly accessible under restricted access. Researchers may request access through the PhysioNet platform after completing institutional ethics review and signing a data use agreement.

## Ethics Approval

Ethical review and approval were obtained from the Institutional Review Boards of Beth Israel Deaconess Medical Center (2001-P-001699/14) and Massachusetts Institute of Technology (No. 0403000206). The study was conducted in accordance with local regulations and institutional policies. As data from MIMIC-IV are anonymized and do not contain any personally identifiable information, the need for written informed consent was waived.

## Author Contributions

Shuyuan Qi: Conceptualization, data curation, formal analysis, writing—original draft preparation.

Yingxiu Huang: Data curation, Methodology.

Shuying Qi: Formal analysis, Manuscript reviewing & editing. Guangyao Zhai: Supervision, Manuscript reviewing & editing.

## Acknowledgment

The authors gratefully acknowledge the valuable contributions of the clinical scientist team throughout the study design and consultation process.

## Supporting information

S1 Table. Univariable analysis for in-hospital mortality and 30-day mortality.

S2 Table. Sensitivity analysis. COX regression models using complete-case analysis (missing data excluded) to assess the association between serum phosphate levels and mortality risks.

## Reference

1. Waksman R, Pahuja M, Van Diepen S, Proudfoot AG, Morrow D, Spitzer E, et al. Standardized Definitions for Cardiogenic Shock Research and Mechanical Circulatory Support Devices: Scientific Expert Panel From the Shock Academic Research Consortium (SHARC). Circulation. 2023;148(14):1113–1126. doi:10.1161/CIRCULATIONAHA.123.064527

2. Osman M, Syed M, Patibandla S, Sulaiman S, Kheiri B, Shah MK, et al. Fifteen-Year Trends in Incidence of Cardiogenic Shock Hospitalization and In-Hospital Mortality in the United States. JAHA. 2021;10(15):e021061. doi:10.1161/JAHA.121.021061

3. Kolte D, Khera S, Aronow WS, Mujib M, Palaniswamy C, Sule S, et al. Trends in Incidence, Management, and Outcomes of Cardiogenic Shock Complicating ST-Elevation Myocardial Infarction in the United States. JAHA. 2014;3(1):e000590. doi:10.1161/JAHA.113.000590

4. Berg DD, Bohula EA, Van Diepen S, Katz JN, Alviar CL, Baird-Zars VM, et al. Epidemiology of Shock in Contemporary Cardiac Intensive Care Units: Data From the Critical Care Cardiology Trials Network Registry. Circ: Cardiovascular Quality and Outcomes. 2019;12(3):e005618. doi:10.1161/CIRCOUTCOMES.119.005618

5. Hunziker L, Radovanovic D, Jeger R, Pedrazzini G, Cuculi F, Urban P, et al. Twenty-Year Trends in the Incidence and Outcome of Cardiogenic Shock in AMIS Plus Registry. Circ: Cardiovascular Interventions. 2019;12(4):e007293. doi:10.1161/CIRCINTERVENTIONS.118.007293

6. Sterling LH, Fernando SM, Talarico R, Qureshi D, van Diepen S, Herridge MS, et al. Long-Term Outcomes of Cardiogenic Shock Complicating Myocardial Infarction. J Am Coll Cardiol. 2023;82(10):985–995. doi:10.1016/j.jacc.2023.06.026

7. Choi KH, Kang D, Park H, Park TK, Lee JM, Song YB, et al. In-hospital and long-term outcomes of cardiogenic shock complicating myocardial infarction versus heart failure. Eur J Heart Fail. 2024;26(7):1594–1603. doi:10.1002/ejhf.3333

8. Thiele H, Zeymer U, Akin I, Behnes M, Rassaf T, Mahabadi AA, et al. Extracorporeal Life Support in Infarct-Related Cardiogenic Shock. N Engl J Med. 2023;389(14):1286–1297. doi:10.1056/NEJMoa2307227

9. Ostadal P, Rokyta R, Karasek J, Kruger A, Vondrakova D, Janotka M, et al. Extracorporeal Membrane Oxygenation in the Therapy of Cardiogenic Shock: Results of the ECMO-CS Randomized Clinical Trial. Circulation. 2023;147(6):454–464. doi:10.1161/CIRCULATIONAHA.122.062949

10. Parlow S, Fernando SM, Pugliese M, Qureshi D, Talarico R, Sterling LH, et al. Resource Utilization and Costs Associated With Cardiogenic Shock Complicating Myocardial Infarction. JACC: Advances. 2024;3(8):101047. doi:10.1016/j.jacadv.2024.101047

11. Kalra S, Ranard LS, Memon S, Rao P, Garan AR, Masoumi A, et al. Risk Prediction in Cardiogenic Shock: Current State of Knowledge, Challenges and Opportunities. Journal of Cardiac Failure. 2021;27(10):1099–1110. doi:10.1016/j.cardfail.2021.08.003

12. Naidu SS, Baran DA, Jentzer JC, Hollenberg SM, Van Diepen S, Basir MB, et al. SCAI SHOCK Stage Classification Expert Consensus Update: A Review and Incorporation of Validation Studies. Journal of the American College of Cardiology. 2022;79(9):933–946. doi:10.1016/j.jacc.2022.01.018

13. Zeymer U, Alushi B, Noc M, Mamas MA, Montalescot G, Fuernau G, et al. Influence of Culprit Lesion Intervention on Outcomes in Infarct-Related Cardiogenic Shock With Cardiac Arrest. Journal of the American College of Cardiology. 2023;81(12):1165–1176. doi:10.1016/j.jacc.2023.01.029

14. Patel SM, Berg DD, Bohula EA, Baird-Zars VM, Barsness GW, Chaudhry SP, et al. Early Serial Assessment of Aggregate Vasoactive Support and Mortality in Cardiogenic Shock: Insights From the Critical Care Cardiology Trials Network Registry. Circ: Heart Failure. 2024;17(5). doi:10.1161/CIRCHEARTFAILURE.124.011736

15. Peacock M. Phosphate Metabolism in Health and Disease. Calcif Tissue Int. 2021;108(1):3–15. doi:10.1007/s00223-020-00686-3

16. Cao W, Li Y, Wen Y, Fang S, Zhao B, Zhang X, et al. Higher serum phosphorus and calcium levels provide prognostic value in patients with acute myocardial infarction. Front Cardiovasc Med. 2022;9:929634. doi:10.3389/fcvm.2022.929634

17. Da J, Xie X, Wolf M, Disthabanchong S, Wang J, Zha Y, et al. Serum Phosphorus and Progression of CKD and Mortality: A Meta-analysis of Cohort Studies. American Journal of Kidney Diseases. 2015;66(2):258–265. doi:10.1053/j.ajkd.2015.01.009

18. Owaki A, Inaguma D, Aoyama I, Inaba S, Koide S, Ito E, et al. Serum phosphate level at initiation of dialysis is associated with all-cause mortality: a multicenter prospective cohort study. Renal Failure. 2018;40(1):475–482. doi:10.1080/0886022X.2018.1499530

19. Li S, Huang Q, Nan W, He B. Association between serum phosphate and in-hospital mortality of patients with AECOPD: A retrospective analysis on eICU database. Heliyon. 2023;9(9):e19748. doi:10.1016/j.heliyon.2023.e19748

20. Nan W, Huang Q, Wan J, Peng Z. Association of serum phosphate and changes in serum phosphate with 28-day mortality in septic shock from MIMIC-IV database. Sci Rep. 2023;13(1):21869. doi:10.1038/s41598-023-49170-6

21. Zhang JF, Jing J, Meng X, Pan Y, Wang YL, Zhao XQ, et al. Serum Phosphate and 1-Year Outcome in Patients With Acute Ischemic Stroke and Transient Ischemic Attack. Front Neurol. 2021;12:652941. doi:10.3389/fneur.2021.652941

22. Kim DW, Jung WJ, Lee DK, Lee KJ, Choi HJ. Association between the initial serum phosphate level and 30-day mortality in blunt trauma patients. J Trauma Acute Care Surg. 2021;91(3):507–513. doi:10.1097/TA.0000000000003271

23. Hedjoudje A, Farha J, Cheurfa C, Grabar S, Weiss E, Badurdeen D, et al. Serum phosphate is associated with mortality among patients admitted to ICU for acute pancreatitis. UEG Journal. 2021;9(5):534–542. doi:10.1002/ueg2.12059

24. Zheng WH, Yao Y, Zhou H, Xu Y, Huang HB. Hyperphosphatemia and Outcomes in Critically Ill Patients: A Systematic Review and Meta-Analysis. Front Med. 2022;9:870637. doi:10.3389/fmed.2022.870637

25. Johnson AEW, Bulgarelli L, Shen L, Gayles A, Shammout A, Horng S, et al. MIMIC-IV, a freely accessible electronic health record dataset. Sci Data. 2023;10(1):1. doi:10.1038/s41597-022-01899-x

26. Ghaferi AA, Schwartz TA, Pawlik TM. STROBE Reporting Guidelines for Observational Studies. JAMA Surg. 2021;156(6):577. doi:10.1001/jamasurg.2021.0528

27. Inker LA, Eneanya ND, Coresh J, Tighiouart H, Wang D, Sang Y, et al. New Creatinine- and Cystatin C–Based Equations to Estimate GFR without Race. N Engl J Med. 2021;385(19):1737–1749. doi:10.1056/NEJMoa2102953

28. Samsky MD, Morrow DA, Proudfoot AG, Hochman JS, Thiele H, Rao SV. Cardiogenic Shock After Acute Myocardial Infarction: A Review. JAMA. 2021;326(18):1840. doi:10.1001/jama.2021.18323

29. Parenica J, Jarkovsky J, Malaska J, Mebazaa A, Gottwaldova J, Helanova K, et al. Infectious Complications and Immune/Inflammatory Response in Cardiogenic Shock Patients: A Prospective Observational Study. Shock. 2017;47(2):165–174. doi:10.1097/SHK.0000000000000756

30. Diakos NA, Thayer K, Swain L, Goud M, Jain P, Kapur NK. Systemic Inflammatory Burden Correlates with Severity and Predicts Outcomes in Patients with Cardiogenic Shock Supported by a Percutaneous Mechanical Assist Device. J of Cardiovasc Trans Res. 2021;14(3):476–483. doi:10.1007/s12265-020-10078-5

31. McCallum W, Testani JM. Updates in Cardiorenal Syndrome. Medical Clinics of North America. 2023;107(4):763–780. doi:10.1016/j.mcna.2023.03.011

32. Chen Y, Luo M, Xu H, Zhao W, He Q. Association between serum phosphate and mortality in critically ill patients: a large retrospective cohort study. BMJ Open. 2021;11(9):e044473. doi:10.1136/bmjopen-2020-044473

33. Turner ME, Beck L, Hill Gallant KM, Chen Y, Moe OW, Kuro-o M, et al. Phosphate in Cardiovascular Disease: From New Insights Into Molecular Mechanisms to Clinical Implications. ATVB. 2024;44(3):584–602. doi:10.1161/ATVBAHA.123.319198

34. Hu W, Jiang S, Liao Y, Li J, Dong F, Guo J, et al. High phosphate impairs arterial endothelial function through AMPK-related pathways in mouse resistance arteries. Acta Physiologica. 2021;231(4):e13595. doi:10.1111/apha.13595

35. Noordali H, Loudon BL, Frenneaux MP, Madhani M. Cardiac metabolism — A promising therapeutic target for heart failure. Pharmacology & Therapeutics. 2018;182:95–114. doi:10.1016/j.pharmthera.2017.08.001

36. Doenst T, Nguyen TD, Abel ED. Cardiac Metabolism in Heart Failure: Implications Beyond ATP Production. Circulation Research. 2013;113(6):709–724. doi:10.1161/CIRCRESAHA.113.300376

37. Reynolds JL, Joannides AJ, Skepper JN, McNair R, Schurgers LJ, Proudfoot D, et al. Human Vascular Smooth Muscle Cells Undergo Vesicle-Mediated Calcification in Response to Changes in Extracellular Calcium and Phosphate Concentrations: A Potential Mechanism for Accelerated Vascular Calcification in ESRD. Journal of the American Society of Nephrology. 2004;15(11):2857–2867. doi:10.1097/01.ASN.0000141960.01035.28

38. Jono S, McKee MD, Murry CE, Shioi A, Nishizawa Y, Mori K, et al. Phosphate Regulation of Vascular Smooth Muscle Cell Calcification. Circulation Research. 2000;87(7). doi:10.1161/01.RES.87.7.e10

39. Guo J, Fujiyoshi A, Willcox B, Choo J, Vishnu A, Hisamatsu T, et al. Increased Aortic Calcification Is Associated With Arterial Stiffness Progression in Multiethnic Middle-Aged Men. Hypertension. 2017;69(1):102–108. doi:10.1161/HYPERTENSIONAHA.116.08459

40. Voelkl J, Egli-Spichtig D, Alesutan I, Wagner CA. Inflammation: a putative link between phosphate metabolism and cardiovascular disease. Clinical Science. 2021;135(1):201–227. doi:10.1042/CS20190895

